# Toward patient-centered tuberculosis preventive treatment: preferences for regimens and formulations in Lima, Peru

**DOI:** 10.1101/2020.06.17.20133942

**Authors:** Courtney M. Yuen, Ana K. Millones, Jerome T. Galea, Daniela Puma, Judith Jimenez, Leonid Lecca, Mercedes C. Becerra, Salmaan Keshavjee

**Affiliations:** Division of Global Health Equity, Brigham and Women’s Hospital, Boston, MA, USA; Department of Global Health and Social Medicine, Harvard Medical School, Boston, MA, USA; Socios En Salud Sucursal Perú, Lima, Peru; School of Social Work, University of South Florida, Tampa, FL, USA; College of Public Health, University of South Florida, Tampa, FL, USA

**Author notes:** Address correspondence to: Courtney M. Yuen, Harvard Medical School, 641 Huntington Avenue, Boston, MA, USA 02115.

## Abstract

**Background:** To ensure patient-centered TB preventive treatment, it is important to consider factors that make it easier for patients to complete treatment. However, there is little published literature about patient preferences for different preventive treatment regimen options, particularly from countries with high tuberculosis burdens.

**Methods:** We conducted a qualitative research study using a framework analysis approach to understand preventive treatment preferences among household contacts. We conducted three focus group discussions with 16 members of families affected by TB in Lima, Peru. Participants were asked to vote for preferred preventive treatment regimens and discuss the reasons behind their choices. Coding followed a deductive approach based on prior research, with data-driven codes added.

**Results:** In total, 7 (44%) participants voted for 3 months isoniazid and rifapentine, 4 (25%) chose 3 months isoniazid and rifampicin, 3 (19%) chose 4 months rifampicin, and 2 (13%) chose 6 months isoniazid. Preferences for shorter regimens over 6 months of isoniazid were driven by concerns over “getting tired” or “getting bored” of taking medications, the difficulty of remembering to take medications, side effects, and interference with daily life. For some, weekly dosing was perceived as being easier to remember and less disruptive, leading to a preference for 3 months isoniazid and rifapentine, which is dosed weekly. However, among caregivers, having a child-friendly formulation was more important than regimen duration. Caregivers reported difficulty in administering pills to children, and preferred treatments available as syrup or dispersible formulations.

**Conclusions:** There is demand for shorter regimens and child-friendly formulations for preventive treatment in a high-burden setting. Individual preferences differ, suggesting that patient-centered care would best be supported by having multiple shorter regimens available.

## INTRODUCTION

TB preventive treatment for contacts is a critical component of an effective response to the global TB epidemic.^1^ However, in settings with high TB burdens, preventive treatment is underutilized.^2^ Adherence is poor for 6 months of daily isoniazid, the only preventive treatment option that has long been available in high-burden settings.^3,4^ However, shorter regimens with equal efficacy are available in high-income countries, and significantly better treatment completion has been observed under programmatic conditions.^5-7^

Much emphasis has recently been placed on creating patient-centered models of TB care, which includes addressing individual patient needs and preferences.^8^ However, there are very few published reports of patients having been asked about their preferences for preventive therapy regimens.^9,10^ Understanding factors that make it easier for patients to finish treatment and offering options that accommodate patient priorities can help programs deliver patient-centered care. Moreover, it is important to acknowledge that preferences may vary among patients. While understanding and accommodating patient preferences may lead to greater complexity in service delivery, it could also maximize treatment success.^11^

We sought to understand the preferred preventive treatment regimens among household contacts of patients being treated for TB in Peru, and the drivers behind these preferences. Given updated World Health Organization guidelines endorsing the expansion of preventive treatment to contacts other than young children,^12^ we wanted to know if answers differed between caregivers considering preventive treatment for their children versus adults considering preventive treatment for themselves. To address these questions, we conducted focus group discussions among people from families affected by TB in Lima, Peru.

## METHODS

We conducted a qualitative research study in Lima, Peru using a framework analysis approach^13^ to analyze focus group discussions on preferences for TB preventive treatment regimens. The objective of the focus groups was to learn which of four WHO-recommended preventive treatment regimens^12^ members of families affected by TB would prefer and to understand the drivers of those preferences. Methods and results are reported according to Consolidated Criteria for Reporting Qualitative Research (COREQ) guidelines.

### Study setting and context

Peru is a middle-income country with an estimated TB incidence of 123 per 100,000 population, placing it among the highest burden TB settings in the Americas.^2^ In Lima, despite widespread awareness of TB disease,^14^ many people lack accurate knowledge about TB.^14, 15^ Moreover, many household contacts of patients with TB are not aware of their increased risk of developing TB,^15^ and use of preventive treatment for contacts is suboptimal.^16-18^

Preventive treatment in Peru is recommended for contacts of patients with drug-susceptible pulmonary TB.^19^ It is explicitly recommended for contacts who are <5 years old or who are 5-19 years old with a positive TST result, and doctors can use their discretion for others who are at high risk. The only currently approved regimen is 6 months of daily isoniazid (6H), which is self-administered. Children who cannot swallow pills are given crushed pills.

We conducted focus group discussions to learn about patient preferences for preventive treatment, including preferences for different regimens and treatment support options; only regimen preferences are presented here. The purpose of the focus groups was to aid the planning and implementation of an intervention to improve contact management, and to provide timely feedback to colleagues involved in the process of revising the national TB guidelines. We considered the possibility that the guideline revision could incorporate regimens other than 6H and could strengthen the recommendation to give preventive treatment to adult contacts. Therefore, we chose to present three shorter WHO-recommended regimens for prevention of drug-sensitive TB to participants,^12^ and to explore preferences for both child and adult contacts.

### Study population

We conducted three focus group discussions in 2019 with members of communities affected by TB in Carabayllo District in Lima, Peru. Participants were purposively sampled from families with a member currently receiving treatment for TB, thus representing the target population for preventive treatment. Participants were recruited either during a treatment support visit to the household or by phone. Focus group 1 comprised adult caregivers aged 18-45 years old; group 2 comprised adults 46-70 years old; and group 3 comprised caregivers of children receiving TB preventive treatment. Groups 1 and 2 were asked about their own preventive treatment preferences while group 3 were asked about the preferences for preventive treatment for their children. No participants dropped out during the discussion.

### Data collection

Focus group discussions were conducted at the Socios En Salud office in Spanish by a female Peruvian nurse (AKM) who was the coordinator for a study evaluating strategies to improve TB contact management in Carabayllo District. In general, the participants did not know the facilitator, but a few had previously met her years ago when she was the coordinator of a program providing support to families affected by TB. In the group 1 discussion, an experienced qualitative researcher (JTG) helped to co-facilitate. In the group 3 discussion, the lead author (CMY) was present as an observer. The facilitator had previously been trained in leading focus group discussions by an experienced Peruvian qualitative researcher.

The facilitator stated to the participants that the goal of the focus group discussions was to understand what families affected by TB want so that TB services can be improved. The facilitator first reviewed basic concepts of TB transmission, latent TB infection, the high risk that contacts have of developing TB, and the purpose of preventive treatment. She then presented WHO-approved preventive treatment regimens for drug-susceptible TB: 6 months of daily isoniazid (6H), 4 months of daily rifampin (4R), 3 months of daily isoniazid and rifampin (3HR), and 3 months of weekly isoniazid and rifapentine (3HP). The facilitator presented a table summarizing the number of pills, dosing frequency, and duration of treatment for each regimen (Table). In addition, isoniazid and rifampin pills were exhibited so that participants could see the size of the pills; a photo was shown of rifapentine pills since rifapentine is not available in Peru. Standard adult doses were presented to groups 1 and 2, and a dose for a 15-20 kg child was presented to the caregivers (group 3). Caregivers were told that 4R was available as a syrup (currently available in Peru) and that 3HR was available as a dispersible tablet that dissolved in water to produce a fruit-flavored liquid (not currently available in Peru).

**Table:** Description of regimens presented to groups 1 and 2

| Option | Description |
| --- | --- |
| A | 1 isoniazid tablet, daily, for 6 months (180 doses) |
| B | 2 rifampin tablets, daily, for 4 months (120 doses) |
| C | 2 rifampin tablets + 1 isoniazid tablet, daily, for 3 months (90 doses) |
| D | 6 rifampin tablets + 3 isoniazid tablets, weekly, for 3 months (12 doses) |

Group 3 was presented a version of this table with dosing for a 15-20 kg child and options for child-friendly formulations

Participants were first asked to indicate the regimen they would choose on an anonymous paper ballot, which was turned into the facilitator. The focus group discussion was then initiated, with the facilitator asking participants to explain the reasons for their preferences. The facilitator used probes such as, “what were the reasons for your choice?”, “which option do you think your family members or neighbors would prefer?”, and “why did you choose this daily treatment instead of this weekly treatment?” (or vice-versa). The discussion of regimen preferences lasted approximately 30 minutes. Focus group discussions were audio recorded and transcribed verbatim by a Peruvian transcription professional who was not a member of the research team. The lead author (CMY) compared the transcripts against the audio recording to assess fidelity; transcripts were not shared with the participants.

### Data analysis

Transcripts were uploaded into the qualitative analysis software Dedoose.^20^ The lead author coded the transcripts in Spanish following a deductive approach based on previous research that had identified regimen characteristics as drivers of preventive treatment regimen preferences among caregivers in Lesotho.^9^ Themes of regimen duration, dosing frequency, and pill burden, and formulation were pre-defined. These pre-defined codes corresponding to regimen characteristics comprised one group in the code tree. In addition, during coding, a separate group of data-derived codes was created corresponding to underlying concerns that caused participants to value certain regimen characteristics. The analysis focused on understanding relationships between these two groups of codes. No participant feedback on coding was solicited.

### Ethical considerations

All participants gave verbal informed consent for participation in the focus group. Transport and compensation for time equivalent to 10 USD was provided. This study was approved by the institutional review boards of Harvard Medical School and the Universidad Peruana Cayetano Heredia.

## RESULTS

### Regimen preferences

The three focus group discussions included 5 younger adults (group 1), 6 older adults (group 2) and 5 caregivers (group 3). Overall, 14 (88%) participants were female. On the ballots, 7 (44%) participants chose 3HP, 4 (25%) chose 3HR, 3 (19%) chose 4R, and 2 (13%) chose 6H. Concerns over medication fatigue, the difficulty of remembering to take medications, side effects, and interference with daily life drove choices for shorter regimens, and in some cases 3HP because of the weekly dosing (Figure). However, among caregivers, the predominant driver of regimen choice was the palatability of the formulation.

**Figure:**
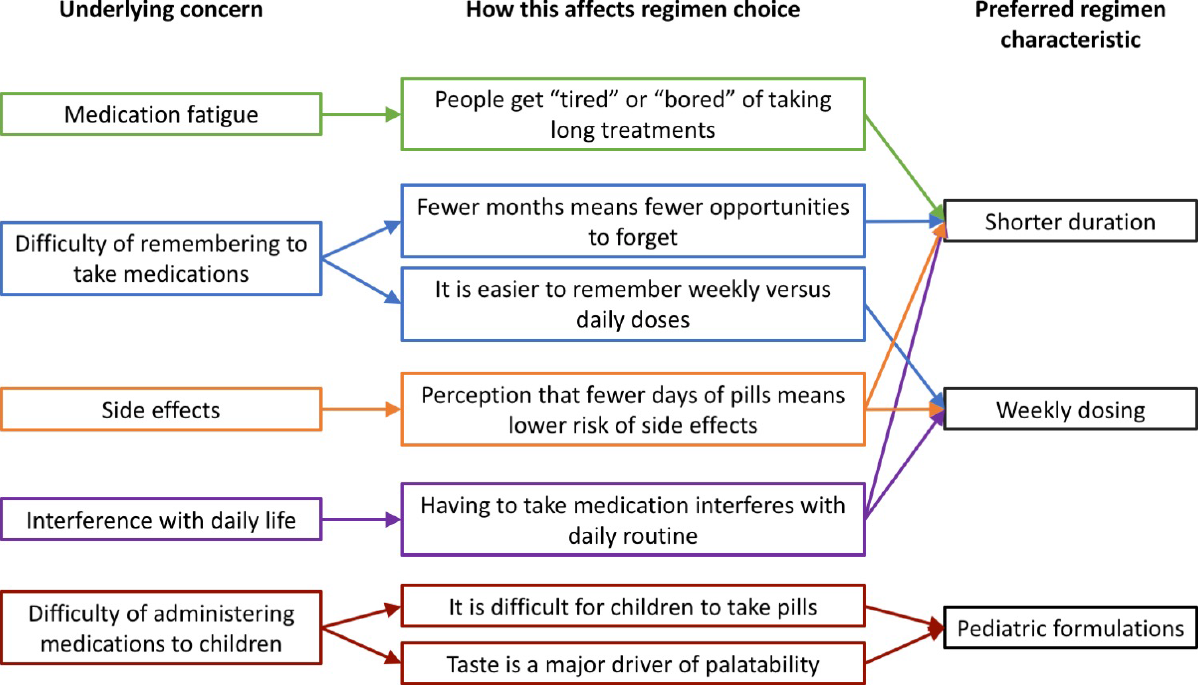
Concerns driving choices of preferred regimens

### Medication fatigue and memory

Participants mentioned the fact that people “get tired” or “get bored” of taking medications over time as a reason for preferring shorter regimens over 6H. Some participants mentioned seeing this medication fatigue occur for their family members. A woman (group 1) commented on her brother-in-law’s experience taking 6H, saying, “At times he gets tired of it and does not want to take it, but he has to continue.” In addition, a male caregiver (group 3) described how his daughter complained to him about the length of time she was on treatment for TB disease.

Memory was also mentioned as a barrier to completing 6H. One woman (group 1) said, “A person will forget, even if it is only once; but in six months they may forget ten times, twenty times – at times, thirty days.” Another woman (group 2) referred to memory as a challenge by recalling a time when she took two doses of her daily medications in a single day because she could not remember if she had taken them or not. Some participants who chose 3HP said that they felt it would be easier to remember to take a weekly dose, such as a woman (group 2) who said, “Because me, I would forget every day. But if it is once a week, I will mark it down and just take it.”

### Side effects

Though the facilitator did not provide participants with any information regarding the side effect profiles of the different regimens, participants in all groups expressed concern about side effects. Minimizing side effects was given as a reason for preferring shorter regimens.

Moreover, participants mentioned side effects when arguing both for and against weekly dosing, depending on whether they felt that taking pills every day or taking more pills at in a single dose would put more stress on the body.

> “My brother-in-law, when he takes [6H], he tells me that it gives him nausea, or maybe [affects] his liver. Well, I would like to take 3 months and no more.” (Woman, group 1)
>
> “There are people with liver problems and taking pills every day will have a greater effect on their fatty liver disease, while taking them once per week would not cause much.” (Woman, group 1)
>
> “It is many pills for his little stomach, and more than anything I consider the health of the child, the little one, that nothing should harm him, and nothing should irritate his stomach.” (Female caregiver, group 3, referring to 3HP)

In addition, one woman (group 2) mentioned that she was taking medications for multiple chronic conditions, and it was important that TB preventive treatment not interact with any of her other medications.

### Interference with daily life

A theme that emerged only in the group 1 (adults 18-45 years old) discussion was the desire to minimize the interference of TB preventive treatment with daily life. Participants expressed the opinion that 3HP would lead to less interference with daily life because of its shorter duration and weekly dosing.

> “Three months compared to six – it is obviously less, right? And it gives you more time to do all your activities, to recuperate.” (Man, group 1)
>
> “Everyone has different routines, everyone has different routines for work, whether it be for studies, whether it be for leisure, or whatever they spend their time doing, right? They would choose [3HP] because if it is once per week…it would prevent them from saying ‘at such a time I have to go do something.’ They would simply say, ‘ah, I have to do this today, and tomorrow no’ and from then, not until the following week.” (Woman, group 1)

### Child-friendly formulations

In the group 3 discussion, all caregivers said they would prefer either 3HR or 4R in a child-friendly formulation, and generally gave reasons related to the formulation as opposed to the duration of treatment. The syrup and dispersible formulations were perceived as being easier to administer than pills. Multiple caregivers mentioned the difficulty they had encountered giving pills to children. Although only one caregiver mentioned familiarity with child-friendly formulations of TB drugs (a male caregiver whose daughter had taken rifampin syrup for TB disease treatment), other caregivers were familiar with child-friendly formulations of other medications. One female caregiver mentioned paracetamol syrup, and another suggested that the dispersible 3HR would be like other effervescent tablets.

> “Partly because of the flavor, and partly because this one is three months [in duration] – for these reasons I chose [3HR], for the taste more than anything. Because I know that children – most children – will take it. I did not pick [3HP]; [3HP] also seems good, a good option, because it is once a week. But the problem seems to me that in the case of children 5 years old and under, they will not take it. If it were for a 10-year-old, yes… well no, it seems like a lot of pills. But [3HR], yes, it seems to me a good choice for children. [4R], no, because the syrup, as they say, has a bad taste. And [6H], two pills, no; children will not swallow them.” (Female caregiver, group 3)

Caregivers mentioned the taste of the medications as a major determinant of whether children would take them. In general, caregivers expected the syrup and dispersible formulations to taste better than crushed or chewed pills. However, the male caregiver, who had tasted his daughter’s rifampin syrup, said that it also did not taste good and that children would not like it. In addition, one female caregiver expressed the concern that if the dispersible tablet is dissolved in a large volume of liquid and the child does not like the taste, the child would not finish the dose.

## DISCUSSION

We found that in Peru, a country with a high TB burden, there is demand for shorter regimens and child-friendly formulations for preventive treatment. The perception that shorter regimens would reduce medication fatigue and interference with daily life suggests that treatment completion could be improved by using shorter regimens, as has been observed in low-burden settings.^5-7^ In addition, we found that caregivers were mainly concerned with having child-friendly formulations. Finally, we observed heterogeneity among participants’ regimen preferences, suggesting that adherence might be best supported by having multiple treatment options available.

We found side effects to be a major concern for participants, in contrast to previous studies of TB preventive treatment preferences. Caregivers in Lesotho interviewed about preventive treatment preferences for their children were not greatly concerned about side effects, as long as they were educated about them.^9^ Moreover, people living with HIV in South Africa ranked side effects as one of their lowest-priority concerns related to preventive treatment, and perceived the benefits as outweighing the risks.^21^ However, a discrete choice experiment in Canada that used hypothetical regimens found that different groups of patients prioritized side effects differently; for some, risk of liver damage was the most important attribute in a hypothetical regimen, while others were more concerned with the clinic visit schedule and the effectiveness of the regimen.^10^ One limitation of our study was that we did not describe regimen-specific side effect profiles to participants and thus do not know whether preferences would have been different if we had. However, our observation that side effect risk was perceived to be related to regimen characteristics such as duration and pill burden highlights the importance of patient education to prevent inaccurate assumptions about side effects.

In our study, child-friendly formulations dominated the discussion among caregivers. Caregivers rejected pill-based regimens and debated between 3HR vs 4R not on the basis of treatment duration but on whether children would take a syrup or a dispersible. This contrasts with the focus on treatment duration and pill burden observed in the aforementioned Lesotho study, in which caregivers were only presented pill-based treatment options.^9^ Notably, 3HP, which was attractive to adult contacts, was rejected by caregivers because of the lack of a child-friendly formulation. We are not aware of studies assessing preferences for child-friendly formulations for TB preventive treatment. However, these preferences have been documented for other medications,^22^ suggesting that preventive treatment completion for child contacts could be improved by making child-friendly formulations available.

While we asked participants to choose a regimen and explain their choice, we did not prompt them with specific regimen attributes and thus cannot comment on the relative importance of different attributes in their decision-making process. For example, the large pill burden in the 3HP dose was mentioned as a barrier in group 3 (caregivers), but not in the other groups. We cannot determine whether this is because group 1 and 2 participants did not care about pill burden or whether it may in fact have been a driver behind the decisions of the group 1 and 2 participants who chose 3HR, 4R, and 6H, but they chose not to articulate it. In addition, despite repeated probing from the facilitator, it was not clear why two participants said that they would prefer 6H, although one woman was taking 6H at the time, so it is possible that familiarity drove her decision. Future studies using discrete choice modeling or conjoint analysis could provide a better understanding about how patients value different regimen attributes and the tradeoffs among them.

Our study had important limitations. We were unable to recruit many men to participate, as men who were approached often declined because of their work schedules. Thus, we were unable to gain insight both into possible sex differences, as well as preferences for adults who work regular schedules outside the home. In addition, because these focus groups were conducted with the main objective of receiving timely programmatic feedback, we were limited to a small number of groups. Although we encountered consistent themes across participants in all three groups, the small sample size limited our ability to capture the full diversity of patient preferences. Finally, while we included in our study caregivers about treatment preferences for their children, we did not include young children, who would be the end-users of child-friendly formulations. Understanding children’s preferences should be included in future studies, since both child and caregiver preferences contribute to a child’s successful treatment completion, and the two may differ.^22^

In conclusion, in a high-burden setting, people in families affected by TB wanted short, easy-to-remember, and non-disruptive TB preventive treatment, with child-friendly formulations for child contacts. Moreover, people expressed different preferences and rationales, meaning that true patient-centered care would involve providing multiple treatment options. Engaging patients to understand their needs and preferences is not only important for building more effective care delivery systems, but also for informing research and development around new treatment options. Current research around even shorter regimens and long-acting formulations could provide even more attractive TB preventive treatment options for patients.^23,24^ However, these new treatments will be most successful in application if they are well matched to patient priorities, including concerns around side effects and ease of administration. To improve TB elimination efforts, it is necessary to make currently available shorter regimens and child-friendly formulations available for TB preventive treatment in high-burden settings, as well as to engage patients during the development of new treatments.

## Data Availability

All data analyzed during this study are included in this published article and its supplementary information files.

## REFERENCES

1. Rangaka MX, Cavalcante SC, Marais BJ, Thim S, Martinson NA, Swaminathan S, et al. Controlling the seedbeds of tuberculosis: diagnosis and treatment of tuberculosis infection. Lancet. 2015;386(10010):2344-53..

2. Global Tuberculosis Report 2019. Geneva: World Health Organization; 2019.

3. Szkwarko D, Hirsch-Moverman Y, Du Plessis L, Du Preez K, Carr C, Mandalakas AM. Child contact management in high tuberculosis burden countries: a mixed-methods systematic review. PLoS One. 2017;12(8):e0182185..

4. Alsdurf H, Hill PC, Matteelli A, Getahun H, Menzies D. The cascade of care in diagnosis and treatment of latent tuberculosis infection: a systematic review and meta-analysis. Lancet Infect Dis. 2016;16(11):1269-78..

5. Cruz AT, Starke JR. Completion rate and safety of tuberculosis infection treatment with shorter regimens. Pediatrics. 2018;141(2)

6. Macaraig MM, Jalees M, Lam C, Burzynski J. Improved treatment completion with shorter treatment regimens for latent tuberculous infection. Int J Tuberc Lung Dis. 2018;22(11):1344-9..

7. Ronald LA, FitzGerald JM, Bartlett-Esquilant G, Schwartzman K, Benedetti A, Boivin JF, et al. Treatment with isoniazid or rifampin for latent tuberculosis infection: population-based study of hepatotoxicity, completion and costs. Eur Respir J. 2020;55(3)

8. Odone A, Roberts B, Dara M, van den Boom M, Kluge H, McKee M. People- and patient-centred care for tuberculosis: models of care for tuberculosis. Int J Tuberc Lung Dis. 2018;22(2):133-8..

9. Hirsch-Moverman Y, Mantell JE, Lebelo L, Wynn C, Hesseling AC, Howard AA, et al. Tuberculosis preventive treatment preferences among care givers of children in Lesotho: a pilot study. Int J Tuberc Lung Dis. 2018;22(8):858-62..

10. Guo N, Marra CA, FitzGerald JM, Elwood RK, Anis AH, Marra F. Patient preference for latent tuberculosis infection preventive treatment: a discrete choice experiment. Value Health. 2011;14(6):937-43..

11. Szkwarko D, Hirsch-Moverman Y. One size does not fit all: preventing tuberculosis among child contacts. BMJ Glob Health. 2019;4(6):e001950..

12. World Health Organization. WHO consolidated guidelines on tuberculosis: module 1: prevention: tuberculosis preventive treatment. Geneva: World Health Organization; 2020.

13. Smith J and Firth J. Qualitative data analysis: the framework approach. Nurse Res. 2011;18(2):52-62..

14. Penaloza R, Navarro JI, Jolly PE, Junkins A, Seas C, Otero L. Health literacy and knowledge related to tuberculosis among outpatients at a referral hospital in Lima, Peru. Res Rep Trop Med. 2019;10:1-10..

15. Shu E, Sobieszczyk ME, Sal Yrvg, Segura P, Galea JT, Lecca L, et al. Knowledge of tuberculosis and vaccine trial preparedness in Lima, Peru. Int J Tuberc Lung Dis. 2017;21(12):1288-93..

16. Yuen CM, Millones AK, Contreras CC, Lecca L, Becerra MC, Keshavjee S. Tuberculosis household accompaniment to improve the contact management cascade: A prospective cohort study. PLoS One. 2019;14(5):e0217104..

17. Otero L, Battaglioli T, Rios J, De la Torre Z, Trocones N, Ordonez C, et al. Contact evaluation and isoniazid preventive therapy among close and household contacts of tuberculosis patients in Lima, Peru: an analysis of routine data. Trop Med Int Health. 2020;25(3):346-56..

18. Chiang SS, Roche S, Contreras C, Del Castillo H, Canales P, Jimenez J, et al. Barriers to the treatment of childhood tuberculous infection and tuberculosis disease: a qualitative study. Int J Tuberc Lung Dis. 2017;21(2):154-60..

19. Ministerio de Salud. Norma técnica de salud para la atención integral de las personas afectadas por tuberculosis. 2013.

20. Dedoose Version 8.0.35, web application for managing, analyzing, and presenting qualitative and mixed method research data. Los Angeles, CA: SocioCultural Research Consultants, LLC; 2018.

21. Kim HY, Hanrahan CF, Dowdy DW, Martinson NA, Golub JE, Bridges JFP. Priorities among HIV-positive individuals for tuberculosis preventive therapies. Int J Tuberc Lung Dis. 2020;24(4):396-402..

22. Ranmal SR, Cram A, Tuleu C. Age-appropriate and acceptable paediatric dosage forms: Insights into end-user perceptions, preferences and practices from the Children’s Acceptability of Oral Formulations (CALF) Study. Int J Pharm. 2016;514(1):296-307..

23. Keshavjee S, Amanullah F, Cattamanchi A, Chaisson R, Dobos KM, Fox GJ, et al. Moving toward tuberculosis elimination. Critical issues for research in diagnostics and therapeutics for tuberculosis infection. Am J Respir Crit Care Med. 2019;199(5):564-71..

24. Swindells S, Siccardi M, Barrett SE, Olsen DB, Grobler JA, Podany AT, et al. Long-acting formulations for the treatment of latent tuberculous infection: opportunities and challenges. Int J Tuberc Lung Dis. 2018;22(2):125-32..

